# Testing the Feasibility of a Smartphone Application for Collecting Data on Occupational Violence Exposure Among Nurses and Midwives

**DOI:** 10.64898/2026.09.14.26363061

**Authors:** Jed Duff, Michael Chataway, Grace Xu, Amanda Fox, Sandra Johnston

**Author notes:** **Corresponding author: Professor Jed Duff**, School of Nursing, Queensland University of Technology, and Royal Brisbane and Women’s Hospital, Australia.

## Abstract

**Background:** Nurses are disproportionately affected by occupational violence, yet conventional incident reporting and retrospective surveys underestimate its extent and miss its context. Ecological momentary assessment (EMA) offers a way to capture violence in near-real time.

**Aim:** To evaluate the feasibility of a smartphone EMA application for capturing the incidence, characteristics, and reporting of occupational violence among hospital nurses and midwives.

**Methods:** In a prospective, single-site feasibility study at a large public hospital, nurses and midwives used a commercially available EMA application (MetricWire) to report occupational violence in two-hourly blocks across their shifts over a three-week period. Incidence, incident characteristics, reporting, and usability were analysed descriptively.

**Findings:** Of 108 enrolled participants, 69 contributed 607 momentary assessments, recording 111 incidents. Violence was reported in 18.6% of assessments, and 62% of participants reported at least one incident. Most incidents were verbal (69%) and patient-perpetrated (86%); 59% involved witnessing violence against a colleague. Although 57% of incidents were reported to someone, only 6% reached the formal incident management system. Respondents (n = 16) found the application easy to use and acceptable.

**Conclusion:** Smartphone-based EMA feasibly and acceptably captures contextually rich, near-real-time data on occupational violence, including the witnessed exposure and under-reporting that conventional systems miss.

**Summary of Relevance:** *Problem or Issue.:* Nurses experience high rates of occupational violence, but conventional incident reporting and retrospective surveys under-capture it and miss its timing, context, and the violence nurses witness against colleagues.

*What is Already Known.:* Occupational violence against nurses is common and harmful. Existing measurement approaches are limited by underreporting and recall bias, and rarely capture incidents in real time or distinguish direct from witnessed exposure.

*What this Paper Adds.:* A smartphone-based ecological momentary assessment application can feasibly and acceptably capture near-real-time, contextually rich data on occupational violence among nurses and midwives. It revealed substantial witnessed exposure (59% of incidents) and a striking gap between incidents reported informally (57%) and those entered into the formal incident-management system (6%).

## Introduction

Occupational violence is a critical issue in healthcare, posing significant threats to the safety, wellbeing, and job satisfaction of healthcare workers, particularly nurses. Defined as any incident in which a person is abused, threatened, or assaulted in circumstances relating to their work, occupational violence can have severe physical, psychological, and economic impacts (Queensland Health, 2016). Its high incidence underscores the urgent need for effective strategies to monitor and mitigate it. This pilot study tests whether a smartphone application can capture near-real-time data on nurses’ and midwives’ exposure to occupational violence in a large Australian public hospital, aiming to overcome the limitations of traditional reporting methods and provide a more comprehensive picture of its incidence and context (Liu et al., 2019; Speroni et al., 2014).

### Background and Rates of Violence Against Nurses

The problem of occupational violence in healthcare is multifaceted. As primary caregivers at the forefront of patient interaction, nurses are particularly susceptible to violent incidents. These range from verbal abuse and threats to physical assault, all of which contribute to a hostile work environment (Gillespie et al., 2010). Such violence affects not only the immediate victim but also the wider healthcare system, contributing to absenteeism, higher attrition, and reduced quality of patient care (Speroni et al., 2014). The psychological toll can lead to long-term mental health problems, including anxiety, depression, and post-traumatic stress disorder (Li et al., 2020).

Occupational violence against nurses is a well-documented global problem. A systematic review estimated that around 62% of healthcare workers had experienced workplace violence (Liu et al., 2019), and studies of nurses report exposure rates as high as 76% (Speroni et al., 2014; Wressell et al., 2018). Perpetrators are most often patients, but also include patients’ relatives and friends and, through horizontal violence, nursing colleagues (Lanctôt & Guay, 2014).

### Contributing Factors of Occupational Violence

The factors contributing to occupational violence are multifaceted, encompassing clinical, organisational, and environmental elements. Clinical factors include patient conditions such as dementia, delirium, mental illness, or head injury, which can heighten aggression (Spelten et al., 2020). In such cases, aggression may itself be a clinical symptom, compounded by situational factors within the hospital (Phillips, 2016). Patients experiencing severe pain, confusion, or frustration may lash out at those caring for them (Camerino et al., 2008), and a history of substance use or psychiatric illness can further increase the likelihood of violence (Gillespie et al., 2010).

Organisational factors also play a substantial role. Inadequate staffing, long waiting times, and poor communication can heighten stress and frustration among patients and families, increasing the likelihood of violence (Campbell et al., 2011; Lim et al., 2022). The physical design of facilities matters too: environments with poor sightlines, isolated areas, or congestion can create opportunities for violence to occur unnoticed (Mayhew & Chappell, 2007).

Environmental factors reflect the broader context of care. High-risk settings, including emergency departments, psychiatric and maternity wards, and geriatric units, tend to see higher rates of violence, owing to high patient volumes, acute presentations, and heightened emotional states (Gates et al., 2011; Hahn et al., 2012). Broader societal attitudes toward healthcare workers and the normalisation of violence in the community may also shape its occurrence in clinical settings.

### Measuring Occupational Violence Against Nurses

Despite its prevalence and consequences, occupational violence remains difficult to measure accurately. Most hospitals rely on retrospective self-report surveys and formal incident-reporting systems, both of which are vulnerable to underreporting and recall bias and provide data only after substantial delay (Speroni et al., 2014; Üzar-Özçetin et al., 2020). Paper-based and legacy electronic reporting systems compound the problem: they are time-consuming and burdensome, and the effort involved discourages nurses from logging incidents (Pompeii et al., 2013). Crucially, because these methods capture incidents in aggregate and after the fact, they obscure the dynamic, context-specific nature of violence, including when, where, and under what circumstances it occurs (Liu et al., 2019).

Underreporting is the central obstacle. Nurses frequently decline to report incidents because they fear retaliation, doubt that reporting will change anything, or have come to regard violence as an inherent part of the job (Spector et al., 2014). Retrospective designs are further weakened by recall bias, as details fade or distort over the days to months between an incident and its documentation (Spector et al., 2014). Reporting also skews toward severe, salient events such as physical assault, while lower-intensity behaviours, such as verbal abuse, intimidation, and incivility, go largely unrecorded. This omission matters: repeated exposure to low-intensity hostility can produce cumulative harm to wellbeing comparable to, or greater than, that of discrete physical assaults (Cortina et al., 2001). An accurate picture of occupational violence therefore depends on capturing the full spectrum of exposure, in close to real time, rather than only its most visible episodes.

### Addressing Measurement Challenges: Ecological Momentary Assessment

Ecological momentary assessment (EMA) offers a way to overcome several of these limitations. EMA collects data in situ through repeated, real-time sampling of participants’ experiences in their natural environments, reducing recall bias and improving ecological validity relative to retrospective surveys (Shiffman et al., 2008; Solymosi et al., 2021). Delivered via smartphone, EMA can capture fine-grained temporal and spatial detail, lower the burden of reporting through simple interfaces, and provide immediate feedback to users (Solymosi et al., 2015). In criminology, smartphone-based EMA has been used to map real-time exposure to crime, disorder, and fear across communities (Brisudová et al., 2024; Chataway et al., 2017), capturing the context of incidents, including location, time, and circumstances, that aggregate reporting systems miss (Solymosi et al., 2021). To date, however, this approach has rarely been applied within workplaces, and almost never to occupational violence in hospitals.

Applying EMA to the hospital setting offers a means of addressing these measurement gaps and generating actionable, near-real-time insight into nurses’ exposure to occupational violence. Accordingly, this pilot study evaluates the feasibility of a commercially available EMA smartphone application for capturing occupational violence among nurses and midwives in a large tertiary hospital, including both direct and witnessed exposure and its immediate context. Specifically, we ask whether the approach can (i) capture the incidence and temporal patterning of occupational violence, and (ii) be implemented acceptably and with adequate engagement in routine clinical practice.

## Methods

### Participants and Setting

We conducted a prospective, single-site feasibility study using smartphone-delivered ecological momentary assessment (EMA). Ethical approval was granted by the relevant Human Research Ethics Committee. Registered and enrolled nurses and midwives who worked at least two days per week at a large public health facility were eligible. The facility employs approximately 4,000 nurses and treats more than one million patients annually. Recruitment and data collection took place over a three-month period, supported by paper flyers distributed across wards. Two members of the research team helped interested staff install the application on their personal smartphones, explained the study, and demonstrated how to activate the surveys. Participants were asked to report any occupational violence in two-hourly blocks across each shift over a three-week period. In total, 108 nurses and midwives enrolled and completed the baseline survey (Table 1); of these, 69 contributed 607 momentary assessments. As an incentive, participants received a $5 coffee voucher.

**Table 1:** Participant demographics (N = 108)

| Characteristic | N (%) |
| --- | --- |
| **Gender** |  |
| Male | 23 (21%) |
| Female | 84 (79%) |
| **Role** |  |
| Student nurse | 14 (13%) |
| Enrolled nurse | 5 (5%) |
| Registered nurse | 81 (76%) |
| Registered midwife | 6 (6%) |
| Nurse practitioner | 1 (1%) |
| **Work status** |  |
| Casual | 5 (5%) |
| Part-time | 15 (14%) |
| Full-time | 73 (68%) |
| Placement | 14 (13%) |
| **Service line** |  |
| Cancer Care Services | 1 (1%) |
| Critical Care and Clinical Support | 38 (41%) |
| Internal Medicine Services | 5 (5%) |
| Mental Health | 5 (5%) |
| Nursing Services | 28 (30%) |
| Surgical and Peri-Op Services | 7 (8%) |
| Women's and Newborn Services | 9 (10%) |
| **Age** (years, M ± SD) | 32 ± 9.67 |
| **Role client-facing** (% , M ± SD) | 93% ± 14.3 |
| **Tenure** (years, M ± SD) | 6 ± 5.7 |
| **Professional experience** (years, M ± SD) | 9 ± 9.14 |

### The Mobile Application

Surveys were delivered through MetricWire (MetricWire Inc.), a commercially available, cloud-based ecological momentary assessment platform with iOS and Android applications. A geofence around the hospital campus triggered surveys when participants were on-site. The home screen was designed to be simple and easy to navigate (Figure 1). Consistent with the approved ethics protocol, participants who reported extreme fear or worry about occupational violence received a targeted email and in-app notification providing contact details for security services and psychological counselling.

**Figure 1:**
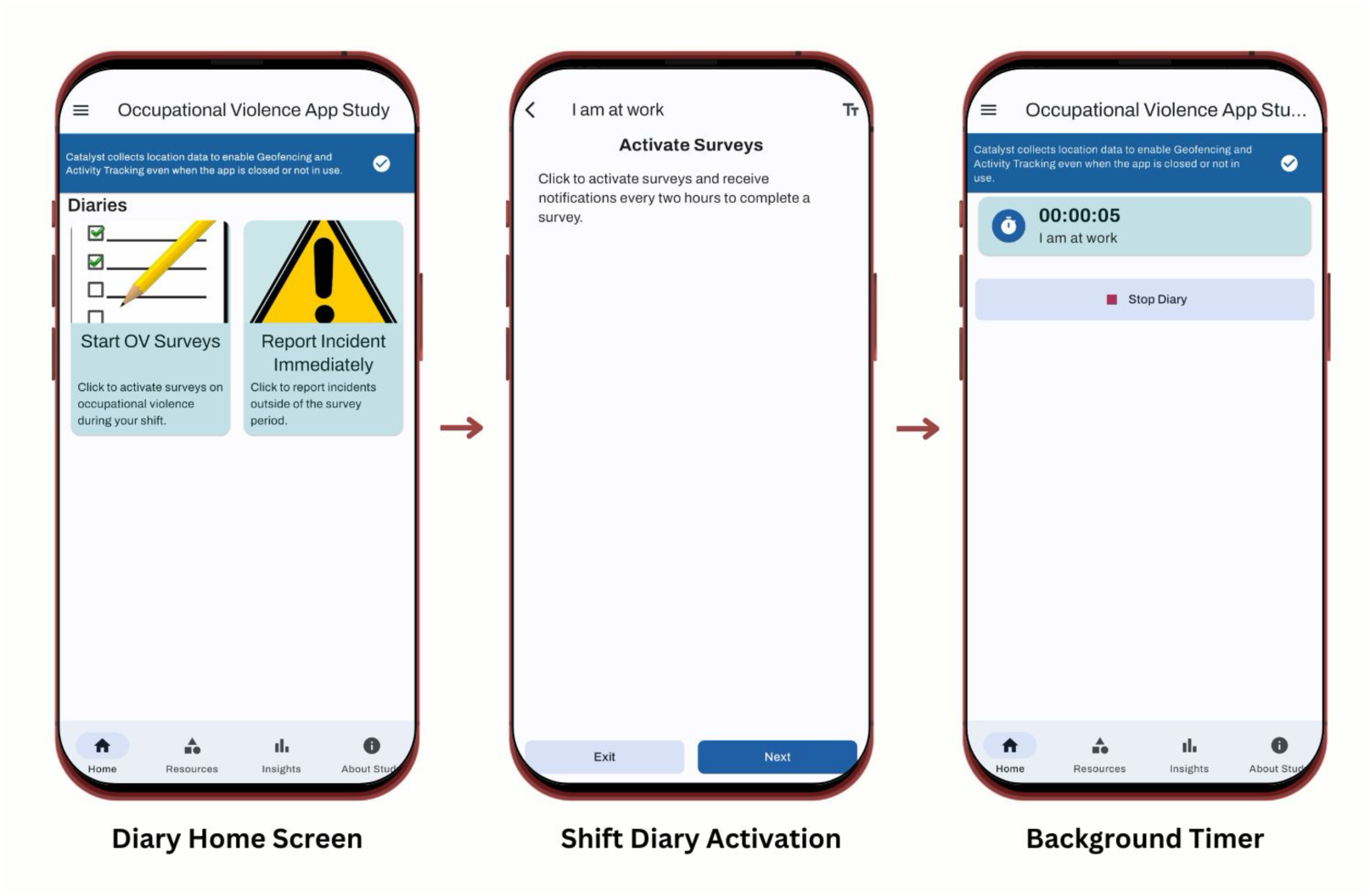
Sample images of the shift-diary activation process in the MetricWire application.

### Instruments

Three instruments were used. A baseline survey (49 items), completed at registration, captured demographic and work characteristics, including ward, role, length of employment, proportion of time in direct patient contact, gender, age, sexuality, and ethnicity (Table 1). The occupational violence EMA was completed during shifts. Every assessment included a brief exposure screen and 4 perceived-risk items (5 items total). When a respondent reported an incident, a further incident module captured its location, timing, duration, perpetrator, exposure type, the respondent’s fear and response (17 items total). Three forms of occupational violence were captured: physical violence (e.g., being hit, slapped, kicked, grabbed, choked, or sexually assaulted), non-physical hostility (e.g., glaring, pacing, photographing or filming staff), and verbal abuse (e.g., swearing, harassment). Every two hours during a shift, participants reported any direct or indirect exposure to these behaviours, with an “I am not at work” option available. At the end of each participant’s three-week period, a brief usability survey (7 items, closed and open-ended) captured perceptions of the app, its acceptability for reporting at work, and the time spent completing surveys.

### Analysis

Analyses were conducted in R and jamovi. Consistent with the design, the unit of analysis was the individual momentary assessment rather than the participant. Incidence was calculated as the number of assessments reporting an incident divided by the number of valid at-work assessments. Temporal patterning was examined descriptively across two-hour blocks and days of the week and displayed as a heatmap (Figure 2); given sparse cell counts, inferential tests of day-by-hour association were not appropriate. Associations between exposure type, violence type, and perpetrator were examined descriptively. Participants’ perceptions of the app were summarised from the usability survey.

**Figure 2:**
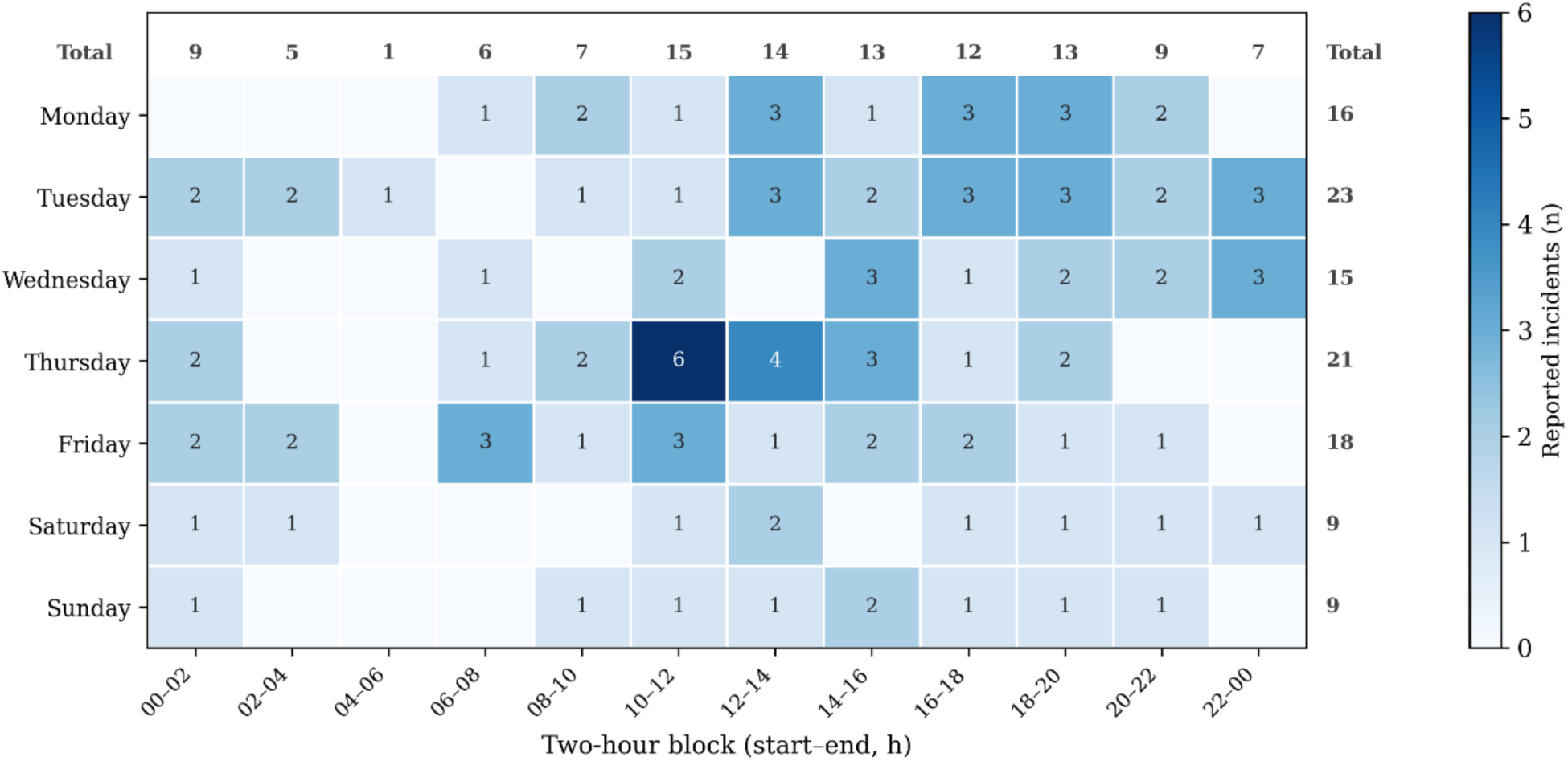
Temporal heatmap of occupational violence reports by day and two-hour block (N = 111).

## Results

### Participant Characteristics

A total of 108 nurses and midwives completed the baseline survey and made up the sample (Table 1). Most were female (84; 79%) and worked full-time (73; 68%). The majority were registered nurses (81; 76%), with smaller numbers of student nurses (14; 13%), registered midwives (6; 6%), enrolled nurses (5; 5%), and one nurse practitioner (1; 1%). Participants were drawn from across the hospital’s service lines, most commonly Critical Care and Clinical Support (38; 41%) and Nursing Services (28; 30%), with the remainder spread across Women’s and Newborn (9; 10%), Surgical and Peri-Operative (7; 8%), Internal Medicine (5; 5%), Mental Health (5; 5%), and Cancer Care (1; 1%) services. On average, participants were 32 years old (SD = 9.67), had 6 years of tenure (SD = 5.7) and 9 years of professional experience (SD = 9.14), and spent 93% of their role in direct patient contact (SD = 14.3).

### Recording the Incidence of Occupational Violence

Of the 108 enrolments, 69 nurses and midwives contributed 607 momentary assessments. These assessments recorded 111 incidents of occupational violence. Pooled across all valid assessments, occupational violence was reported in 18.6% of assessments, approximately one in five. At the participant level, 43 of the 69 contributing nurses and midwives (62%) reported at least one incident during the study. Table 2 presents the number of incidents and the incidence rate within each two-hour block.

**Table 2:** Occupational violence incidence by two-hour block (597 at-work assessments; 111 incidents)

| Two-hour block | Incidents | Valid assessments | Incidence (%) |
| --- | --- | --- | --- |
| 00:00–02:00 | 9 | 26 | 35 |
| 02:00–04:00 | 5 | 17 | 29 |
| 04:00–06:00 | 1 | 8 | 12 |
| 06:00–08:00 | 6 | 36 | 17 |
| 08:00–10:00 | 7 | 61 | 11 |
| 10:00–12:00 | 15 | 94 | 16 |
| 12:00–14:00 | 14 | 89 | 16 |
| 14:00–16:00 | 13 | 73 | 18 |
| 16:00–18:00 | 12 | 70 | 17 |
| 18:00–20:00 | 13 | 58 | 22 |
| 20:00–22:00 | 9 | 46 | 20 |
| 22:00–00:00 | 7 | 19 | 37 |
| **Total** | **111** | **597** | **18.6** |

### Temporal Patterns of Occupational Violence

Incident reports were distributed across the day and week (Table 2). More incidents were recorded on weekdays than weekends (93 vs 18), and more across the late-morning-to-evening period than overnight, with the highest single block being 10:00–12:00 (15 incidents). Incidence rates per assessment were more uniform, ranging from 11% to 37% across two-hour blocks, with the highest rates occurring in the sparsely sampled overnight periods. Given the small number of incidents and the uneven number of assessments across periods, these temporal patterns are presented graphically (Figure 2).

### Perpetrator and Incident Characteristics

Across the 111 incident reports, verbal abuse was the most common form of occupational violence, recorded in 77 incidents (69%), followed by non-physical hostility (47; 42%) and physical violence (20; 18%); these categories were not mutually exclusive, as a single incident could involve more than one form. The person responsible was most often a patient or client: 95 incidents (86%) involved a patient or client alone, and a patient or client was involved in 98 incidents (88%) overall. The remaining incidents involved a relative or friend of the patient, a member of the public, or another individual.

The perpetrator profile differed by the form of violence. All 20 physical violence incidents were perpetrated by a patient or client, and verbal abuse was likewise overwhelmingly patient-perpetrated (96% of verbal incidents). Non-physical hostility, while still most often involving a patient or client (74%), more frequently involved a relative, friend, or visitor (26%) than any other form. Given the small number of incidents and sparse cell counts, these patterns are reported descriptively rather than tested inferentially.

Exposure was frequently both direct and indirect. Nurses were the direct target in 74 incidents (67%) and reported witnessing violence directed at a colleague in 65 incidents (59%); 32 incidents (29%) involved both. That well over half of incidents involved witnessing violence toward a colleague underscores the importance of capturing indirect exposure, which conventional incident reporting typically omits.

### Incident Reporting

Although most incidents were acknowledged in some way, few entered the formal record. Of the 111 incidents, 63 (57%) had been reported to at least one person, most commonly a manager or team leader (39; 35%), security (29; 26%), or a colleague (27; 24%); four were reported to police. By contrast, only 7 incidents (6%) had been entered into the hospital’s formal incident-management system. Among incidents that were not formally reported, the most frequently cited reasons were being too busy or short of time (13), regarding the incident as unimportant (13), and believing that nothing would be done (11).

### Compliance and Engagement

We assessed the feasibility of the EMA approach using data from participants’ devices and the usability survey. The response rate to survey notifications was 58%, with participants completing just over half of the prompts delivered over their three-week period. This represents moderate engagement with the notification schedule. However, where surveys were started, item-level completion was very high at 98%, indicating that the surveys themselves were quick to complete once opened.

### Perceptions of the Application

Sixteen nurses and midwives completed the usability survey. The application was rated easy to use (M = 8.7/10, SD = 1.6) and, on average, acceptable for reporting occupational violence at work (M = 8.4/10, SD = 2.2). Participants rated the time each survey took as neither too little nor too much (M = 5.6/10, SD = 1.6).

Two usability issues recurred. First, several participants reported battery drain; one nurse noted that “leaving the app running causes battery drain”. Second, three nurses preferred multiple-choice questions to the sliding scales, and one found the frequency of notifications too high; one participant suggested an additional end-of-shift survey to capture ongoing perceptions of risk and worry. In free-text feedback, one nurse felt the automated support messages were too frequent and could be limited to incidents involving extreme fear or worry.

## Discussion

The purpose of this pilot study was to test the feasibility of a smartphone application for collecting near-real-time data on nurses’ and midwives’ exposure to occupational violence in a large Australian public hospital. Our findings indicate that the approach is feasible and can substantially improve the quality of occupational violence data, capturing not only the incidence but also its timing, type, perpetrator, and the nature of exposure. We discuss the key findings below and outline directions for extending this methodology across hospital and community care settings.

### Enhancing Data on the Incidence of Occupational Violence

This study captured a substantial burden of occupational violence. Over the three-week period, 62% of participating nurses and midwives reported at least one incident, and violence was recorded in approximately one in five completed assessments (18.6%). Although the short observation window makes direct comparison difficult, this aligns with the high prevalence reported in the wider literature: Spector et al. (2014) found that 36.4% of nurses experience physical violence at work, and Liu et al. (2019) reported that around 62% encounter some form of workplace violence annually. Capturing this burden in near-real time and in context, recording the timing, type, perpetrator, and nature of each incident, offers a more granular picture than annual recall surveys provide.

Retrospective and incident-report-based methods are widely recognised to underestimate the true rate of occupational violence, owing to recall bias and underreporting (Arnetz et al., 2015). Our data illustrate this gap directly. While 57% of incidents captured by the application had been reported to someone, most often a manager (35%), security (26%), or a colleague (24%), only 6% had been entered into the hospital’s formal incident-management system. The great majority of incidents that staff acknowledged, and often escalated informally, never reached the official record on which workforce-safety decisions depend. Where reasons were given for not reporting, nurses most often cited being too busy, regarding the incident as unimportant, or believing that nothing would be done, echoing well-documented barriers to reporting (Spector et al., 2014). By lowering the time and effort required to record an incident, smartphone-based EMA may both improve the completeness of incident data and surface the substantial volume of violence that current systems miss. Better unit-level data has, in turn, been shown to enable effective prevention: in a randomised controlled trial, feeding unit-specific violence data back to hospital teams to guide action planning significantly reduced patient-to-worker violence and related injury (Arnetz et al., 2017).

A further contribution of this approach is its ability to distinguish between types of exposure. Much healthcare research measures only direct victimisation, overlooking indirect exposure (Liu et al., 2019; Spector et al., 2014). This is a meaningful omission: witnessing a colleague being abused or assaulted can produce psychological harm, including anxiety, distress, and reduced wellbeing, comparable to that of direct victimisation (Nielsen & Einarsen, 2018; Tehrani, 2004). In this study, well over half of incidents (59%) involved witnessing violence directed at a colleague, and nearly a third (29%) involved both direct and indirect exposure. Capturing this fuller picture is essential for estimating the true burden of occupational violence and for designing responses that support bystanders as well as direct victims.

These findings have practical implications for hospital incident-monitoring systems. Reporting mechanisms that capture all forms and directions of exposure would give health services a more accurate measure of occupational violence, supporting targeted prevention and a stronger safety culture (Schat & Kelloway, 2006). Accurate documentation also serves a legal function: under Australian work health and safety legislation, for example the Work Health and Safety Act 2011, employers must keep records of workplace incidents and hazards, with regulators monitoring compliance. Smartphone-based reporting offers a practical mechanism for meeting these obligations while improving an organisation’s capacity to detect, monitor, and respond to occupational violence.

### The Temporal Patterning of Occupational Violence

The application also captured the timing of incidents, information that aggregate incident reports rarely provide. Descriptively, more incidents were recorded on weekdays than weekends (93 vs 18) and across the late-morning-to-evening period than overnight. This pattern must be interpreted cautiously, however. Because survey completion was itself concentrated in these periods, the raw counts largely track when participants were active in the study rather than when risk was greatest. Expressed as a rate per completed assessment, incidence was relatively stable across the day and, if anything, marginally higher at weekends (21% vs 18% on weekdays) and during the sparsely sampled overnight blocks, where estimates are imprecise.

This distinction illustrates a key advantage of EMA over count-based incident reporting: by capturing the denominator of exposure, it guards against mistaking patterns of activity for patterns of risk. It also tempers any direct reading of these data against the criminological literature, in which crime and calls for service typically peak on weekends and in the evening (Weisburd, 2015). Several mechanisms could plausibly drive genuine temporal variation in hospital violence, including rostering and shift structure, patient acuity and activity, and visitor presence (Liu et al., 2019), but this pilot cannot separate these from sampling effects. Establishing real temporal patterns, and any resulting implications for staffing or security, will require larger samples with stratified sampling across shifts and days and, where ethically feasible, linkage to rostered hours.

### The Perceived Usability of the Application

Participants found the application a workable way to record occupational violence at the point of care. It was rated easy to use, and most considered it an acceptable means of reporting violence at work, although acceptability ratings were more variable than ratings of ease. This broad usability is consistent with criminological studies that have used smartphone EMA to collect real-time data on victimisation and fear of crime in community samples (Chataway et al., 2017; Engström & Kronkvist, 2022; Kronkvist, 2022; Solymosi et al., 2015).

The most common usability issue was battery drain, reported by several participants. This most likely reflects the geofencing and location sensing used to trigger on-site prompts, although older devices and inconsistent charging may also have contributed. Future deployments could mitigate this by limiting location sensing to rostered hours or deactivating it when a device is idle or off-site, by providing clearer charging guidance, or by supplying battery packs. A second issue concerned response format: several nurses found the visual analogue sliding scales, used to capture reactions such as fear and worry, difficult to use, preferring discrete options. The optimal response format for momentary affect items is debated; in a recent experimental comparison, Haslbeck et al. (2025) found that visual analogue scales may be preferable to Likert scales for capturing affective states related to general psychopathology, particularly where variation occurs near the scale limits. Given the brevity required of EMA items and the need to minimise respondent burden, future occupational-violence EMA studies should continue to test how response format affects data quality and participant experience.

### Limitations

Several limitations should be noted. First, we used a convenience sample, weighted toward nurses working in the emergency department. This limits generalisability across nursing specialties and constrains our ability to examine associations between occupational violence and individual characteristics such as gender, age, and tenure. Future studies would benefit from stratified or quota sampling to obtain a more representative sample.

Second, compliance and engagement were moderate, and reporting was unevenly distributed: of the 108 nurses and midwives who enrolled, 69 contributed momentary assessments, and a small number of participants accounted for a disproportionate share of these. This skew, together with the modest number of incidents, limited the analyses we could undertake and the precision of incidence estimates, particularly within sparsely sampled periods. Improving compliance would support more sophisticated modelling that accounts for the nested structure of EMA data. Strategies for doing so are debated: incentives can improve adherence but may reduce data quality and raise concerns about coercion among vulnerable groups (Kleiman et al., 2017; Porras-Segovia et al., 2020). Communicating the purpose and benefits of the research, and incorporating engagement features such as personal feedback, may encourage reporting without relying solely on incentives (Chataway, 2020).

Third, the instrument captured only three categories of violence (physical, verbal, and non-physical hostility) perpetrated by patients, relatives, or visitors. While this kept the EMA brief, it may oversimplify the nature of occupational violence and, notably, excludes horizontal violence perpetrated by colleagues, which nurses commonly report. Future instruments should consider a broader set of categories, including co-worker–perpetrated violence.

## Future Directions

This study demonstrates the feasibility of using smartphone-based EMA to collect near-real-time data on occupational violence among nurses and midwives, and points to several avenues for further research. First, the approach could be extended to examine the factors contributing to incidents. As outlined earlier, occupational violence has clinical (e.g., pain, psychopathology), environmental (e.g., wait times, patient demand), and organisational (e.g., communication) antecedents. Because EMA captures incidents in context, it could help establish whether particular factors are associated with violence at specific times or in specific areas of a hospital, supporting more targeted prevention. Establishing any temporal or spatial patterning reliably will, however, require the larger, stratified samples noted above.

Second, the approach should be tested with other healthcare workers. Although nurses are disproportionately affected, occupational violence is experienced across clinical and non-clinical hospital roles, and the method may transfer to other settings (e.g., aged care, community care, general practice) and to other high-risk industries such as retail, hospitality, security, and transport.

Third, future work should explore integrating incident capture with real-time responses, such as alerts that mobilise support for affected staff or security assistance. Aggregated, real-time dashboards could also help organisations monitor and anticipate high-risk periods (Jia et al., 2020), enabling them not only to improve the quality of incident data but to respond proactively to emerging risks.

## Conclusion

Occupational violence is a serious and persistent threat to the health and safety of nurses, yet the systems used to detect and record it remain inadequate. This study demonstrates that a smartphone-based EMA application can feasibly and acceptably capture near-real-time data on nurses’ exposure to occupational violence in an acute hospital setting. Beyond counting incidents, the approach captured their timing, type, and perpetrator and, importantly, both direct and witnessed exposure, while revealing how few incidents reach formal reporting systems. In doing so, it addresses a clear gap in how occupational violence is currently measured. With further development and testing in larger and more diverse samples, this approach has the potential to improve the way occupational violence is detected, monitored, and ultimately prevented across healthcare settings.

## Ethics

This project received ethical clearance from the Queensland Health Metro North Human Research Ethics Committee (HREC/2022/MNHA/87412).

## Author contributions (CRediT)

Jed Duff - Conceptualisation, Methodology, Formal analysis, Writing (original draft), Writing (review & editing). Michael Chataway - Conceptualisation, Methodology, Formal analysis, Writing (original draft), Writing (review & editing). Grace Xu - Conceptualisation, Writing (review & editing). Amanda Fox - Conceptualisation, Writing (review & editing). Sandra Johnston - Conceptualisation, Writing (review & editing).

## Data availability

The data that support the findings of this study are available from the corresponding author upon reasonable request.

## Declaration of Generative AI and AI-Assisted Technologies in the Writing Process

During the preparation of this work, the authors used Claude (Anthropic) to assist with language editing, restructuring, and improving the clarity of the manuscript. After using this tool, the authors reviewed and edited the content as needed and take full responsibility for the content of the publication.

